# An artificial intelligence-powered digital pathology platform to support large-scale deworming programs against soil-transmitted helminthiasis and intestinal schistosomiasis in resource-limited settings

**DOI:** 10.1101/2025.08.07.25332931

**Authors:** Peter K. Ward, Mohammed Aliy Mohammed, Mio Ayana, Lindsay A. Broadfield, Peter Dahlberg, Daniel Dana, Gemechu Leta, Zeleke Mekonnen, Betty Nabatte, Narcis Kabatereine, Kristina M. Orrling, Sofie Van Hoecke, Bruno Levecke, Lieven J. Stuyver

## Abstract

**Background:** The World Health Organization (WHO) has emphasised the need for innovative diagnostic tools to support the control and eliminate neglected tropical diseases (NTDs). Microscopy-based diagnostics, the current standard, rely on trained technicians for labour-intensive processes, posing logistical challenges in the low-resource settings where NTDs are most prevalent. This study describes the technical details of an artificial intelligence-powered digital pathology (AI-DP) platform designed to support large-scale deworming programs for two NTDs, alongside its analytical performance and user experience in laboratory and field settings.

**Methodology/Principal Findings:** The AI-DP platform integrates whole slide imaging scanners, onboard AI analysis, electronic data capture tools, and result verification software to automate microscopy-based screening. Targeting soil-transmitted helminths (STH) and intestinal schistosomiasis (SCH) as initial use cases, the system was deployed in Ethiopia and Uganda, scanning 951 Kato-Katz (KK) slides containing 43,919 verified helminth eggs. Using 5-fold cross-validation, precision/recall/average precision were 95.4/91.7/97.1% for *Ascaris lumbricoides*, 95.9/86.7/94.8% for *Trichuris trichiura*, 84.6/86.6/91.4% for hookworm, and 89.1/79.1/89.2% for *Schistosoma mansoni*. Feedback from 14 field users across 30 real-world scenarios noted the AI-DP platform’s improved usability, particularly in hardware portability and software interfaces, though the average scan time of 12.5 minutes per slide was identified as a limitation compared to manual microscopy.

**Conclusions/Significance:** The AI-DP platform demonstrates potential as a tool for efficient monitoring and evaluation of STH and SCH control programs by providing near-real-time data with quality controls. However, further validation studies are needed to assess clinical diagnostic performance, field usability, and cost-effectiveness in large-scale STH and SCH deworming programs. A platform approach supports scalability to other microscopy-based diagnostics, aligning with global elimination goals for NTDs.

**AUTHOR SUMMARY:** Neglected tropical diseases such as soil-transmitted helminthiasis (STH) and schistosomiasis (SCH) afflict over a billion people in low-resource areas, yet diagnosis often depends on labour-intensive manual microscopy performed by trained technicians. We developed a portable, artificial intelligence–powered digital pathology (AI-DP) platform to automate this process, incorporating a field-deployable slide scanner, onboard AI for egg detection and classification, and a user-friendly verification interface for technicians to review results. Designed with consideration of World Health Organization diagnostic needs for NTDs, the platform was tested in laboratory and zero-infrastructure field trials in Ethiopia and Uganda, it processed nearly 1,000 slides, detecting STH and SCH with over 90% precision and recall. Field users noted improvements in portability and ease of use, though scan times remain slower than manual methods. While the AI-DP platform shows potential as a tool for efficient monitoring and evaluation of STH and SCH control programs, with possible extension to other diseases, further validation studies are essential to evaluate its clinical diagnostic performance and cost-effectiveness in large-scale deworming initiatives.

## 1 INTRODUCTION

Neglected tropical diseases (NTDs) are a diverse group of parasitic, bacterial, viral and toxin-mediated conditions that collectively affect over 1.5 billion people [1]. These diseases disproportionately impact vulnerable populations in (sub)tropical countries with limited access to medical care and undermine both individual and community development. The World Health Organization (WHO) has identified the lack of accurate, reliable, affordable diagnostics as a major barrier to achieving the 2030 targets for NTD control and elimination [1]. To guide the development of new tools, WHO has published 18 target product profiles (TPPs) covering 13 NTDs, each outlining desired performance characteristics for future diagnostics [2].

Microscopy remains the mostly widely used method for diagnosing NTDs [1], including both soil-transmitted helminthiasis (STH; infections by *Ascaris lumbricoides*, *Trichuris trichiura*, and hookworms (*Necator americanus* and *Ancylostoma* spp.) and schistosomiasis (SCH, infections by *Schistosoma mansoni* (intestinal SCH) and *S. haematobium* (urinary SCH)), which affect an estimated 1.5 billion people and 251 million people, respectively [3,4]. For STH and intestinal SCH, the WHO-recommended Kato-Katz (KK) method is widely used to prepare stool samples for visual egg counting, providing data on both prevalence and infection intensity [5]. Although the KK method is inexpensive, simple, and rapid, it suffers from low sensitivity at light infection intensities, rapid degeneration of hookworm eggs (within one hour), and the requirement for trained technicians [1,6–8].

Beyond these technical limitations, KK-based surveys incur substantial personnel costs, accounting for 42–74 % of total study expenses [9]. Moreover, personnel requirements scale with sample throughput and the use of duplicate or triplicate smears per participant, making labour the single largest cost component in STH and SCH monitoring programs [9–11]. As programs shift from morbidity control toward elimination as a public health problem, the need for higher-coverage surveys, rising salaries, and dwindling skilled capacity will further escalate operational costs [10–12]. In recognition of these challenges, WHO has emphasised the urgent need for improved diagnostics and workflows in its STH and SCH TPPs [13,14].

Given the limited prospects for non-stool-based methods by 2030 [12], automating aspects of microscopy to boost throughput and reduce technician burden represents a practical strategy. Recent efforts have focused on developing low-cost, automated optical devices integrated with artificial intelligence—hereafter referred to as artificial intelligence–powered digital pathology (AI-DP)—to support monitoring and evaluation (M&E) of STH and SCH control programs [15–17]. These approaches leverage advanced imaging hardware and machine-learning algorithms with the aim to enhance diagnostic accuracy, scalability, and cost-effectiveness by reducing reliance on skilled personnel and accelerating data acquisition. However, these efforts still highlight the numerous technical, operational and regulatory hurdles to address WHO TPP criteria[15].

For example, our initial proof-of-concept AI-DP platform [18] suffered from low throughput, bulky mains-only hardware, inconsistent focus (exacerbated by variable smear thickness and slide quality), and labour-intensive, error-prone ground-truth annotation processes.

This study aims (i) to describe the technical details of the AI-DP platform that was designed based on the aforementioned lessons learned, to report on (ii) the analytical performance of the AI model when using the updated platform and (iii) the user experience when used in both a laboratory and field setting. Note that a formal protocol for clinical diagnostic performance evaluation and usability of this AI-DP platform has been published [19] and will be reported separately once results are available.

## 2 METHODS

### 2.1 Ethics Statement

The study was approved by the Jimma University Institutional Review Board (IHRPGD/555/21) in Ethiopia and by the Ugandan Ministry of Health Vector Control Division Research Ethics Committee (VCDREC154) for both the Integrated Community-based Survey for Program Monitoring (ICSPM) and the precision mapping surveys for SCH. Written informed consent was obtained from all adult participants and from parents or legal guardians of minors; children aged ≥12 years in Ethiopia and ≥8 years in Uganda provided written assent. Participant confidentiality was maintained via pseudonymised QR coded identifiers, and all data were encrypted in transit. Individuals diagnosed positive by manual KK were treated per national guidelines: single-dose albendazole (400 mg) for STH and praziquantel (40 mg/kg) for intestinal SCH.

### 2.2 AI-DP Platform Design

The platform was designed according to our AI-DP specific interpretation of the WHO TPPs for STH [20], with emphasis on the M&E of as a use case. In consultation with the AI4NTD consortium partners, we defined data workflows and product requirements to guide the platform’s design and development. Based on this consultation, the workflow of the AI-DP platform now consists of three major steps (**Fig 1**). In a **first step**, metadata of participant, sample and study site is captured via a cross-platform mobile and web application that handles participant registration (electronic data capture (EDC), quick response (QR) coded sample tracking, and offline caching, with secure synchronisation as soon as connectivity is restored.

**Fig 1.**
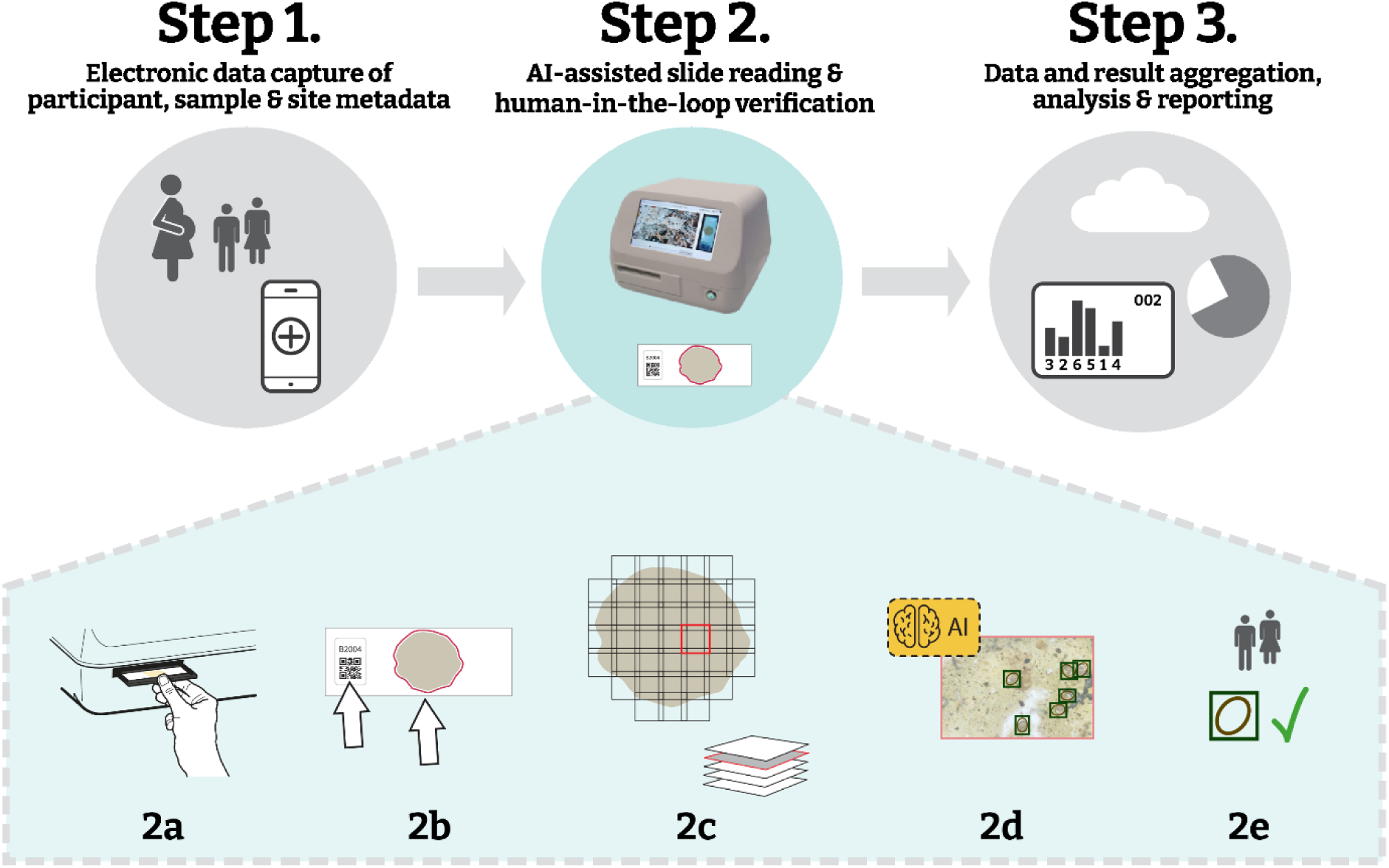
Overview of the AI-powered digital pathology (AI-DP) workflow. **Step 1**, electronic data capture of participant, sample and site metadata using a mobile app; **Step 2**, AI-assisted slide reading with human-in-the-loop verification, comprising **(2a)** loading a standard glass slide into the scanner, **(2b)** QR-code readout and automated detection of scan area, **(2c)** automated focus-stack acquisition across the specimen, **(2d)** onboard AI detection of helminth eggs, and **(2e)** human verification of AI annotations; and **Step 3**, aggregation, analysis and reporting of results via a cloud-connected dashboard.

In the **second step**, KK slides are automatically scanned and helminth eggs are counted with human-in-loop-verification. Generally, this step consists of key operations with the whole slide imaging (WSI) scanner: a barcode-labelled slide is loaded into the cradle of the WSI scanner (step **2a**), the QR code is read and the smear boundaries are mapped (step **2b**), and the device captures focus-stacked image tiles across all fields of view (step **2c**). Each tile stack is submitted to our trained convolutional neural network for egg detection and classification (step **2d**), and the resulting annotations are presented in the EggInspector interface for rapid technician confirmation or correction (step **2e**). This human-in-the-loop verification ensures high sensitivity and specificity before results are transmitted to the reporting dashboard or exported for downstream analysis.

In the **third step**, a cloud-accessible reporting dashboard aggregates technician-verified egg counts alongside survey metadata, offers interactive visualisations of infection prevalence and intensity, and supports both export of results and direct integration with national health information systems.

To cover both extremes of M&E survey settings, we designed the AI-DP platform for school-based surveys in ‘zero-infrastructure’ locations having no grid power, running water, or internet, and laboratory-based surveys with intermittent access to utilities. We packed all necessary hardware into a single ruggedised case (gross weight <32 kg; see **Fig 2**), including (i) two portable WSI scanners, (ii) a Slide Manager server (local image repository and AI inference), (iii) a dedicated laptop for study management and human-in-loop verification of AI detections, (iv) uninterruptible power supply (UPS) with sealed lead-acid battery, (v) label printer, (vi) slide holders, (vii) power and data cables, (viii) WiFi router for high-bandwidth local networking and (ix) printed user manuals. The case with foam inlay prevents the equipment from vibration-induced misalignment and damage during air travel or rough-road transport.

**Fig 2.**
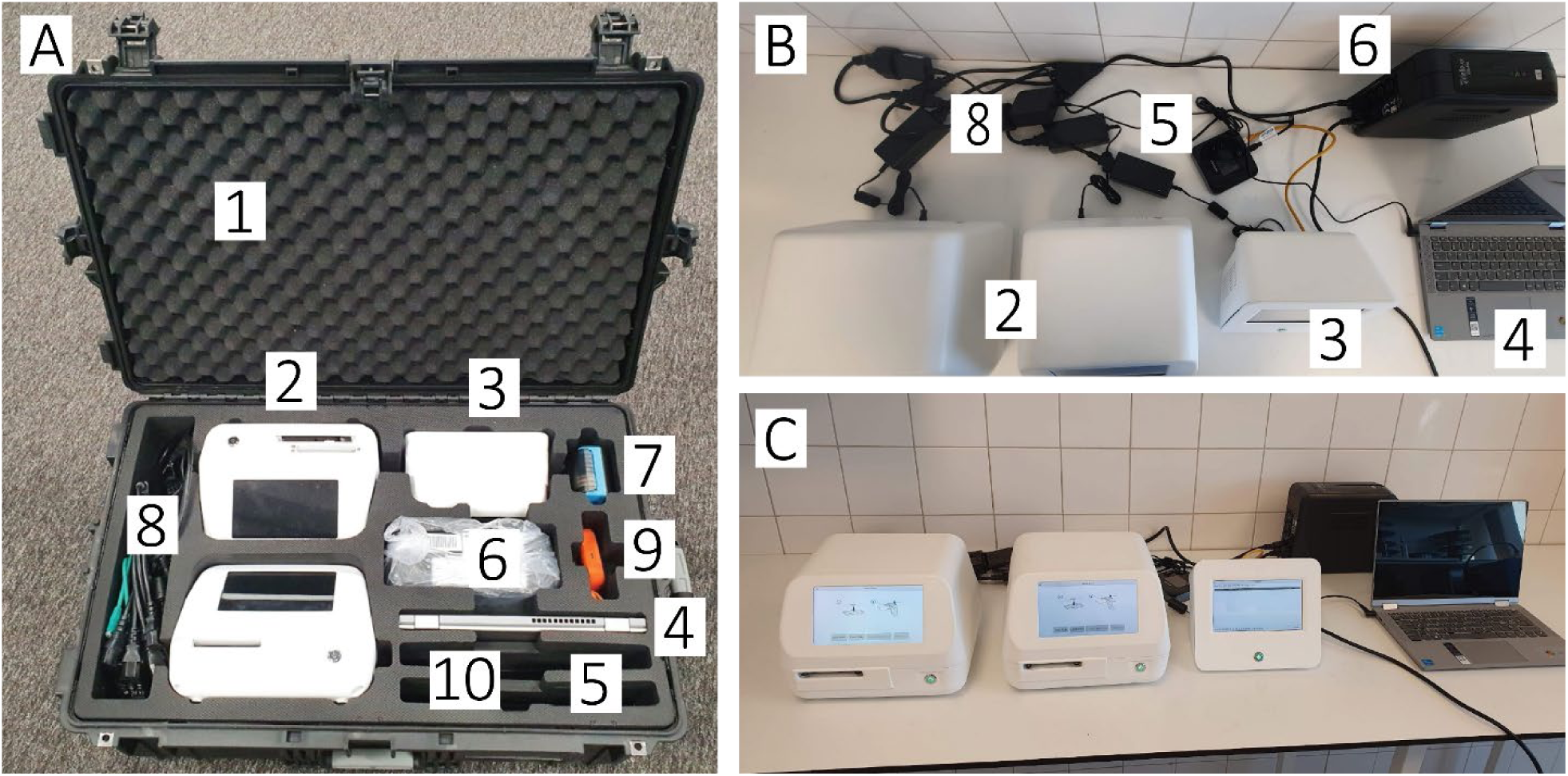
Field-deployable kit for the AI-powered digital-pathology (AI-DP) platform. **(A)** Contents of the ruggedised transport case (total weight <32 kg), showing: (1) protective case with custom foam insert; (2) two whole-slide imaging (WSI) scanners; (3) slide-manager unit for local storage (up to 1 000 scans) and AI processing; (4) laptop computer; (5) Wi-Fi router; (6) uninterruptible power supply (UPS); (7) label printer; (8) network and power cables; (9) 4 TB USB external hard drive; (10) user manuals. **(B)** Top view of unpacked equipment with all connections: the two WSI scanners (2), slide manager (3), laptop (4), Wi-Fi router (5), UPS (6) and power/ethernet cables (8). **(C)** Fully assembled system on the laboratory bench, ready for slide digitisation, data capture and on-site AI inference.

In the following sections we will provide more technical details on the most important components of the AI-DP platform.

#### 2.2.1 Electronic Data Capture

To enable a fully digital, end-to-end workflow, the AI-DP platform assigns a unique QR code label to every sample container, slide, and result form. A mobile EDC app scans these labels (Fig 3) and automatically links each sample identifier to timestamps (collection, preparation, and slide-reading), site registration details and participant demographics, geospatial coordinates, and optional water, sanitation, and hygiene (WASH) indicators. Data can be captured offline, synchronised to a central or cloud-based repository, and exchanged with established EDC systems—such as Open Data Kit (ODK) and District Health Information System 2 (DHIS2)—via application programming interfaces (APIs), ensuring interoperability with national health information systems.

**Fig 3.**
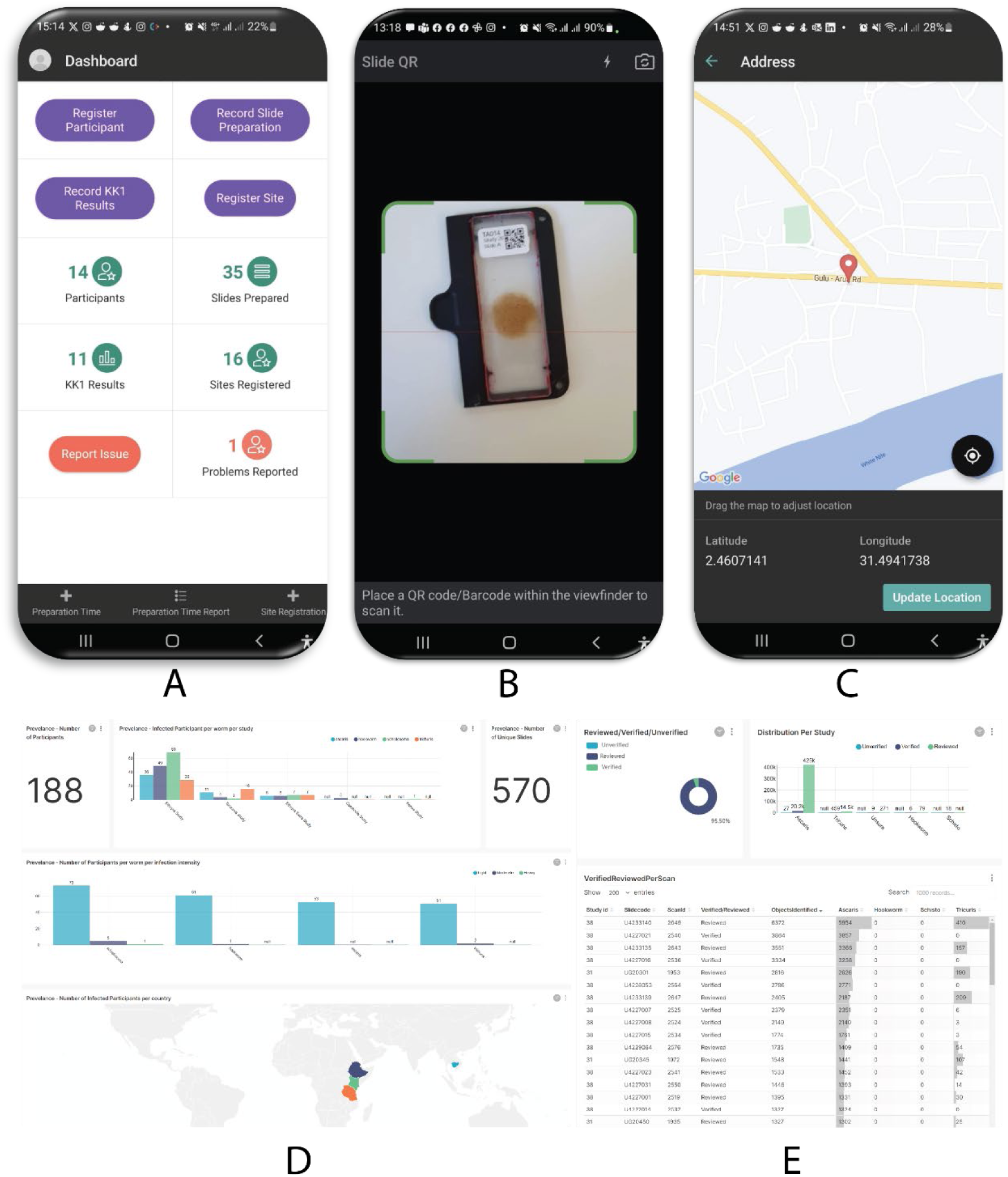
Electronic data capture (EDC) application and reporting dashboard. (A) Customisable main screen of the mobile EDC app for registering participants and sites, and collecting metadata such as human-microscopy Kato-Katz egg counts (KK1). (B) Built-in QR-code scanner to timestamp events such as slide-preparation. (C) Geolocation interface for mapping sample-collection sites, with adjustable pin and latitude/longitude display. (D) Web-based dashboard presenting real-time data summaries, prevalence charts and overall study-progress metrics. (E) Detailed verification panel showing per-scan status (verified, reviewed, unverified) alongside egg-count results in tabular form.

#### 2.2.2 Whole Slide Image Scanner

We designed the custom WSI scanner for two-button operation (‘Load slide’ and ‘Start scan’). Standard 75 × 25 mm glass slides are clamped into a magnetic holder on a motorised XY stage. A secondary overview camera reads each slide’s QR code, confirms the sample type and outline of the KK thick smear, and presents the scan region to the user for confirmation. Operators may adjust manually and start the proceed to start the scan. During scanning, the stage advances the slide through the region while a high-resolution camera captures images at every field of view. The WSI scanners were powered by direct current (DC) 19 V from common laptop chargers for continuous operation, or off a 19V 27,000 mAh USB power bank for up to 3 hours. The WSI scanners included a 1 TB drive permitting approximately 1,000 scanned slides. On-board AI inference runs on an NVIDIA Jetson Xavier NX in a standalone setup or on a NVIDIA Jetson Nano in a high throughput scenario when paired with a Slide Manager (discussed further under AI development).

Imaging is performed with a 1/2.6-inch, 2-megapixel CMOS sensor (3×3 μm pixels) coupled to an inverted lens (7.5 mm focal length, 32.5 mm back focal length) yielding 3.2× magnification, a 1,280 × 720 μm field of view, a single pixel resolution of 0.93 μm, and a theoretical optical resolution of 1.2 μm. This provides an optical resolution is comparable to traditional brightfield microscopy with a 10×/0.25NA Plan Achromat objective. The camera streams at 120 frames per second with illumination being provided by an LED source and diffuser. Captured images may be digitally enlarged via the verification tool.

KK thick smears typically span 20 mm in diameter and 135 μm in thickness (from 41.7 mg stool) with local thickness variations. To ensure at least one in-focus image per field of view, the scanner captures Z-stacks of 8 images at 20 μm intervals (total depth 240 μm) using a liquid lens (having ±0.67 mm range and 2.8 μm step size). A Sobel-filter focus metric ranks each stack; the single sharpest image is processed by the AI model for helminth egg detection and classification. Users may also perform manual refocusing post-scan, analogous to standard microscope workflows.

#### 2.2.3 AI Result Generation

Image processing runs either on-board each WSI scanner in standalone mode or centrally on a Slide Manager when multiple scanners are deployed. The Slide Manager—powered by an NVIDIA Jetson AGX Xavier—provides substantially more compute than the Jetson Nano modules in individual scanners. In centralised operation, images from up to five scanners stream wirelessly to the Slide Manager, which performs AI inference, consolidates storage, and delivers real-time results. This architecture simplifies data management, quality control, and remote verification. During development tests, five scanners ran in parallel under one Slide Manager, while in field deployments in Uganda and Ethiopia, we routinely paired two scanners with a single Manager.

During scanning, each field of view image is sent to the Slide Manager, where an offline AI model automatically detects, classifies, and quantifies helminth eggs as images are acquired. We followed the KK protocol to convert raw egg counts to eggs per gram (EPG) [5]. Field of view images include a 10% overlap (≈100 µm) in both axes to ensure full smear coverage and reduce edge artifacts. To prevent double-counting, we grouped duplicate detections across overlapping frames. We then categorised EPG values as negative, light, moderate, or heavy infection according to WHO thresholds [23]. Per species results are available immediately in the local interface and, when internet connectivity is available, can be uploaded—optionally, with accompanying images—to a cloud-based dashboard for further analysis and remote verification.

#### 2.2.4 Result Verification and Data Annotation Tool

To support diagnostic validation and continuous AI improvement, we developed EggInspector, a human-in-the-loop verification and annotation application available in both offline (local field) and online (cloud-synchronised) modes. On launch (**Fig 4A**), users choose between local data or previously synchronised studies. The Browse tab (**Fig 4B**) lists scanned slides, their fields of view, and detection filters (by species or verification status). In the Verify Objects view (**Fig 4C**), a slide overview shows AI detection overlays alongside a species-specific list of candidate eggs. Verification proceeds in a 3×3 grid of cropped thumbnails (**Fig 4D**), one species at a time; users may accept, reject, or reclassify each detection. Selecting any thumbnail opens the full field of view image (**Fig 4E**), highlights all detections, and enables focal-plane adjustment via the captured z-stack.

**Fig 4.**
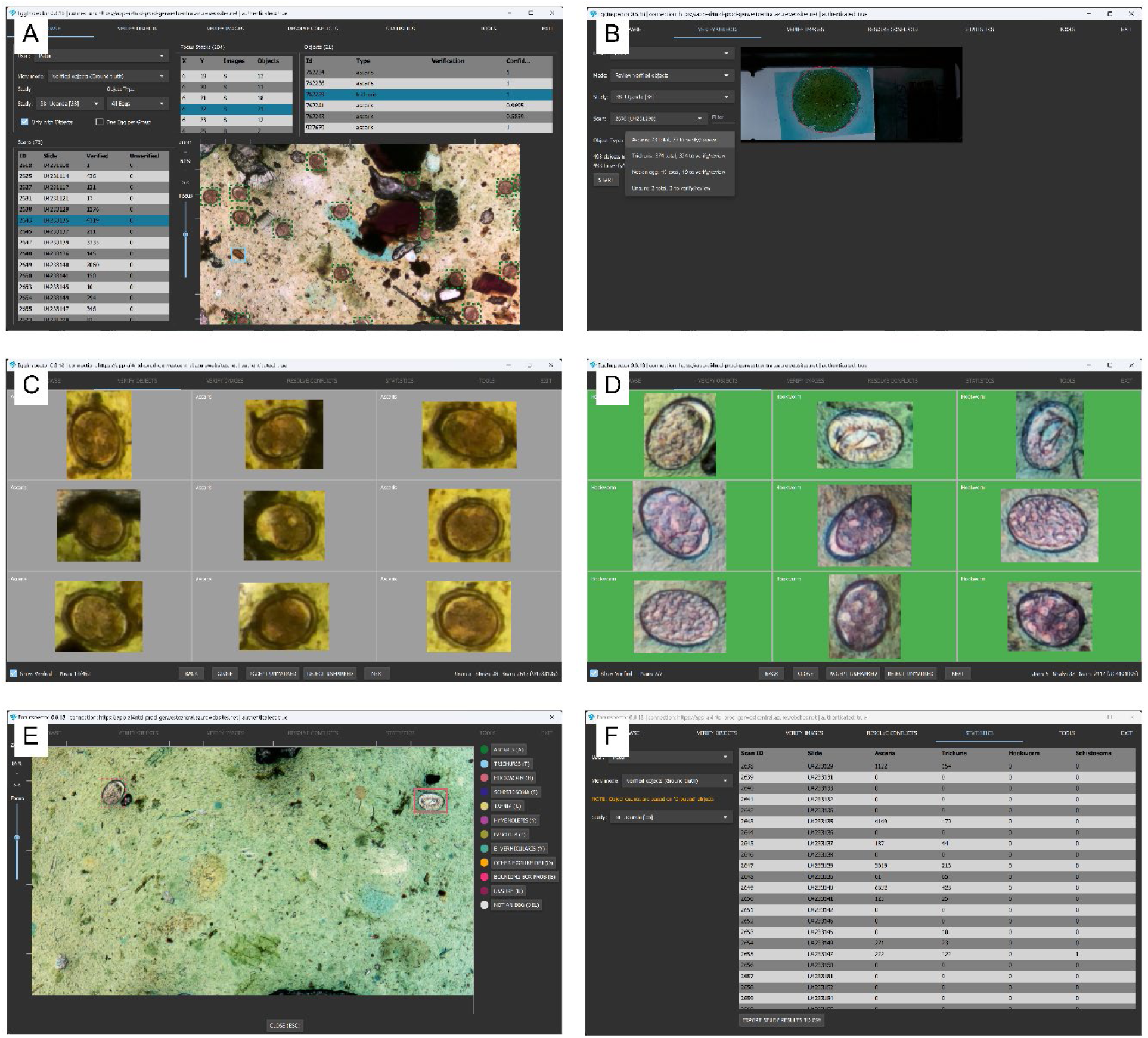
EggInspector, a human-in-the-loop AI verification and image annotation tool. **(A) ‘**Browse’ page lists scanned slides with selectable fields of view, detection-status filters (by species or verification status) and table of candidate detections. **(B)** ‘Verify Objects’ page displaying a selected scan with options to filter objects for verification, and a summary of the verification status. **(C)** Batch verification grid (3×3 cropped thumbnails) of unverified *Ascaris lumbricoides* eggs, with Accept/Reject/Reclassify controls. **(D)** Batch verification grid of verified hookworm eggs. **(E)** Contextual view of a selected hookworm egg within the full field of view, with all other detected eggs highlighted and controls for z-stack focus adjustment and image adjustments. **(F)** Summary page reporting total egg counts by species, eggs per gram (EPG) and options to export results locally or via the central dashboard.

Once verification is complete, EggInspector aggregates species-level egg counts and exports a summary for local storage or cloud upload (**Fig 4F**). This hybrid AI-technician workflow improves diagnostic sensitivity by leveraging optimised AI models and provides specificity by allowing a user to confirm positive detections and catch false positive detections. Optional manual annotation of missed eggs during model-training sessions can further enhance recall.

#### 2.2.5 Data Verification and Reporting

After technician verification via EggInspector, we take the exported egg counts and convert them to EPG using the standard KK protocol [5]. We then assign light, moderate, or heavy infection categories per species using WHO thresholds [21]. Verified results—including slide-level EPG values, associated images, and metadata—are stored locally and, when internet access is available, automatically synchronised to a secure, cloud-based dashboard. This setup enables near-real-time visualisation of study progress, facilitates secondary remote verification by expert reviewers, and supports integration with national health information systems. In low-connectivity settings, completed scan datasets can optionally be exported to external storage devices (e.g., USB drives) and uploaded to central servers once the connection is restored.

### 2.3 Analytical Performance of Artificial Intelligence

The analytical performance of our AI model was assessed through four stages: scanning of slides, ground truth dataset creation, model training and selection, and statistical data analysis.

#### 2.3.1 Scanning of KK slides

Stool samples were collected in two NTD endemic countries (Ethiopia and Uganda). In Ethiopia, we recruited students aged 5–15 years (balanced by sex) from eleven primary schools in Jimma town. In Uganda, we enrolled two cohorts—schoolchildren (5–14 years) and community members aged ≥6 months—in the West Central and West Nile regions. Each participant provided one stool sample in a labelled container. In Jimma, samples were transported at ambient temperature to the university laboratory; in Uganda, preparation, microscopy, and slide scanning were performed onsite under zero-infrastructure conditions (typically no running water, electricity, or internet).

Stool specimens were processed using the WHO KK method [5]. In brief, approximately 41.7 mg of sieved stool was transferred through a standard template onto a glass slide, covered with a glycerol–malachite green–soaked cellophane strip, and pressed flat. Each slide received a preprinted QR label encoding slide ID, sample ID, study ID, date, and time for automated tracking by the WSI scanner.

For AI training and evaluation, we only scanned positive KK slides confirmed by conventional microscopy of either STH or intestinal SCH. Slides were normally scanned 30–60 minutes after preparation to capture hookworm eggs before degeneration. When manual microscopy took priority, some slides were scanned up to 24 hours later to for improved *S. mansoni* egg visibility [5].

#### 2.3.2 Ground Truth Dataset Creation

We assembled a high-quality WSI dataset with image tiles organised by slide ID. Each image record included slide and image IDs, species class labels and bounding box coordinates (x_min, y_min, x_max, y_max). Annotation proceeded in three phases to ensure label accuracy: (i) eight parasitologists independently drew bounding boxes and assigned species labels in EggInspector; (ii) the same parasitologists then provided a second review for images they had not previously seen during their first round of annotation. During this review, verifiers could do one of the following:

- accept the image, confirming all visible labels and acknowledging that no visible eggs were missing;
- change visible objects to an ‘Unsure’ label if uncertain, reject individual objects if confident they were not helminth eggs,
- reassign a label to another helminth type (e.g., *Hymenolepis*, *Taenia*, *Fasciola*, or an ‘Other’ category for unspecified helminths), while identifying and annotating any omissions.

In case of a disagreements, (iii) a third parasitologist adjudicated any disagreements to reach a consensus by reviewing only the objects in conflict. Here too, the adjudicator could accept, reject, mark the object as unsure, or change the label type. Images with unresolved conflicts, meaning no majority consensus was achieved on the object label even after review by multiple verifiers (e.g., differing opinions on species classification or whether an object is an egg), were excluded from the ground truth dataset until a conflict resolution process, such as a majority vote on the label, could be completed. Only images verified by at least two parasitologists with agreement on the labelled objects were retained for training, ensuring reliability and consistency in the dataset.

#### 2.3.3 Model Training and Selection

To prevent data leakage, we employed a grouped five-fold cross-validation strategy (implemented with scikit-learn’s GroupKFold [22]), ensuring all images from the same slide were assigned to a single fold. Each fold was trained on four partitions and evaluated on the held-out partition. We applied transfer learning with the pretrained YOLOv8n model (COCO weights) [23], selecting the nano variant for its compact size and fast inference capabilities on embedded hardware. Training was initialised for 100 epochs per fold on a Tesla T4 graphics processing unit (GPU, 15 GiB) using Python 3.10.13 and PyTorch 2.1.2, using the default Ultralytics YOLOv8 hyperparameters with early stopping to prevent overfitting. The epoch with the highest Ultralytics evaluation score was selected as the best model for each fold. All code and datasets are publicly available on Kaggle [24,25].

#### 2.3.4 Statistical Analysis

For each fold and each helminth species, we computed the following metrics at the object level: Precision (positive predictive value), defined as

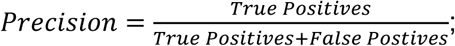

Recall (sensitivity or true positive rate), defined as

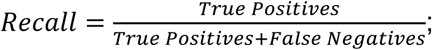

Average precision at an intersection over union of 0.50 (AP50), defined as the area under the precision–recall curve at a 50% overlap threshold [26].

Due to the design of our dataset, which exclusively targeted slides confirmed positive for STH or SCH infections to maximise annotation of helminth eggs for initial model training, we lacked negative-control slides (i.e., slides confirmed free of helminth eggs) necessary to compute diagnostic specificity or negative predictive value. While object-level precision, recall and AP50 characterise the model’s per-egg detection performance, slide-level specificity will be assessed in future work incorporating egg-free specimens.

### 2.4 User Experience

To evaluate the user experience of the platform, informal user feedback, operational observations and written feedback were gathered from users.

## 3 RESULTS

### 3.1 AI Performance

We scanned 951 KK slides in this study (Uganda: n = 620; Ethiopia: n = 331), producing 1,156 whole slide images (Uganda: 636; Ethiopia: 520). Slides were included only if conventional microscopy confirmed the presence of eggs. From these scans, we extracted 8,695 field of view images that each contained at least one egg. In total, 43,919 egg annotations were verified independently by two parasitologists. Species-level distribution was as follows: *A. lumbricoides* (38,854 eggs; 88.5%), *T. trichiura* (3,220 eggs; 7.3%), hookworm (548 eggs; 1.2%), and *S. mansoni* (1,297 eggs; 3.0%). This curated, expert-verified dataset formed the ground truth for model training and cross-validation.

We evaluated detection and classification performance using five-fold GroupKFold cross-validation with slide-based grouping, as detailed in the Methods section. **Table 1** reports the mean ± standard deviation (SD) of precision, recall, and AP50 across the five folds; the full fold-wise breakdown is provided in **S1 Table**.

**Table 1.**
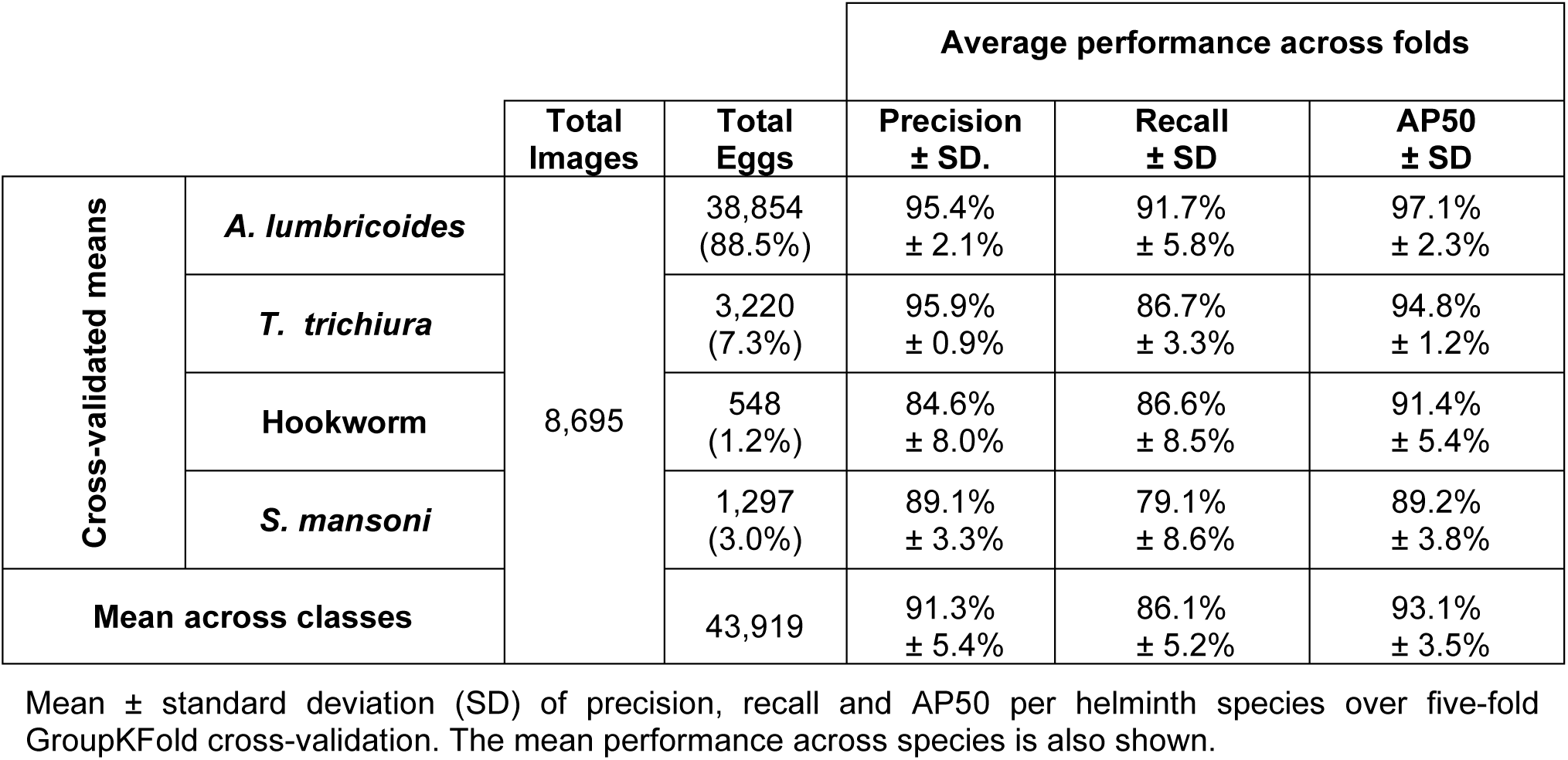
Summary of AI model performance with k-fold cross-validation.

While all species exceed 84% precision and 79% recall, *A. lumbricoides* and *T. trichiura* perform best across all three metrics. Hookworm has the lowest precision (84.6%) and a lower AP50 (91.4%). *S. mansoni* shows the lowest recall (79.1%) and AP50 (89.2%). Both classes are underrepresented in our dataset, suggesting that additional negative- and positive-sample augmentation for these eggs could further boost model performance. For field deployment, we retrained the YOLOv8n model on the full annotated dataset for 70 epochs (the average of the optimal epochs from each cross-validation fold). The frozen model was exported in ONNX format and distributed to edge devices via over-the-air updates. Benchmarking on 1,000 sample images yielded mean inference times of 240 ms/image on NVIDIA Jetson AGX Xavier, 234 ms/image on NVIDIA Jetson Xavier NX, 770 ms/image on NVIDIA Jetson Nano. These results demonstrated that the platform could perform near–real-time analysis even on resource-constrained hardware.

### 3.2 User Experience

During the development of the platform, several versions of the system were evaluated by 14 users across two countries and a range of diverse settings. These settings included standard laboratory environments, simulated field conditions (**Fig 5A**), and over 30 real-world field scenarios (**Fig 5B-D**).

**Fig 5.** The artificial intelligence-powered digital pathology (AI-DP) platform was tested in various field settings and conditions. **(A)** Simulated field testing and system training in Belgium, by members from Jimma University and Ghent University. Additional testing was performed in laboratory environments at both Jimma University and Ghent University. **(B-D)** The AI-DP platform was transported to and evaluated in over 30 zero-infrastructure field sites, across six districts throughout Uganda by the Vector Control Division, Ministry of Health, Uganda, to perform data collection, evaluate the system in remote field settings, test digital workflows, and test integrated AI and data verification workflows. At each site slides were read first by traditional microscopy to screen for positive samples which were then read by the AI-DP platform to support rapid build-up of the database for AI training.

The user observations/feedback and design recommendations for the AI-DP platform are summarised in **Table 2**. We grouped the observations feedback and the corresponding recommendations into five themes, including (i) hardware usability, (ii) power and portability, (iii) scanning performance, (iv) software and interface and (v) training and support. Generally, the AI-DP platform demonstrated a high degree of adaptability operating effectively in both laboratory environments and remote, off-grid locations.

**Table 2.**
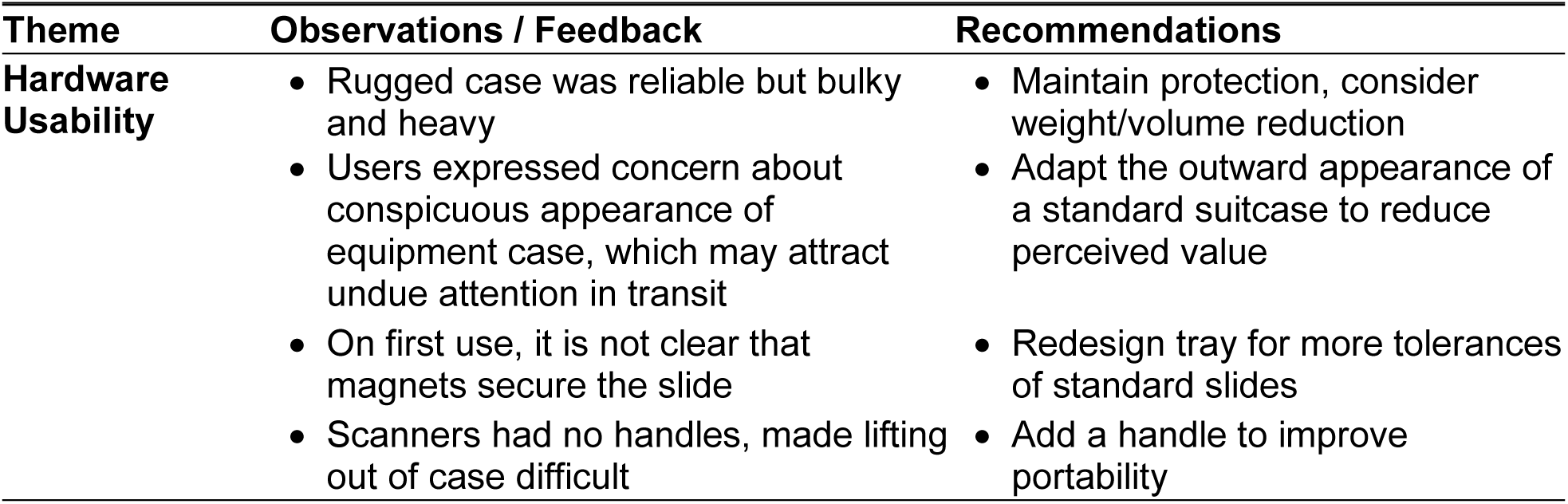
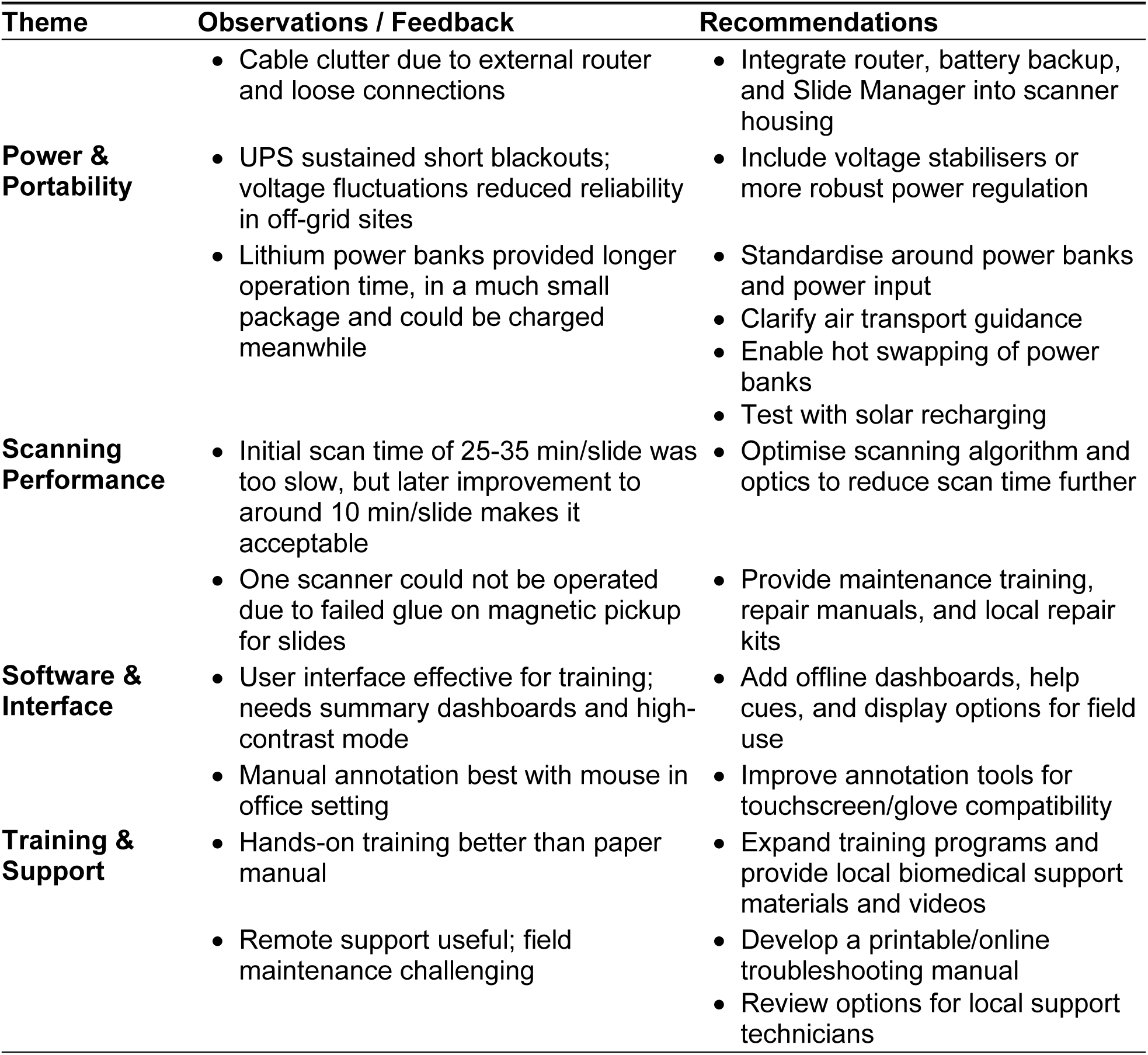
Summary of User Feedback and Design Recommendations for the AI-DP Platform.

#### 3.2.1 Hardware Usability

Although users considered the equipment transport case bulky, it provided robust protection for the equipment during air and ground transport. Fieldwork in Uganda required long daily commutes over bumpy and challenging dirt roads to reach various schools, where the equipment was set up and studies were conducted. Users expressed concern about the conspicuous appearance of the case, noting it could attract undue attention during transit. Additionally, on first use, it was unclear to some users that magnets secured the slide in the scanner tray, leading to potential handling issues. The lack of handles on the scanners made lifting them out of the case difficult, hindering setup efficiency. Lastly, cable clutter from an external router and loose connections was reported as a usability challenge in field settings. Recommendations to address these issues include reducing the case’s weight and volume while maintaining protection, adapting its outward appearance to resemble a standard suitcase to lower perceived value, redesigning the tray for greater tolerance of standard slides, adding handles to improve scanner portability, and integrating the router, battery backup, and Slide Manager into the scanner housing to minimise cable clutter.

#### 3.2.2 Power & Portability

In laboratory settings, the UPS could sustain operations during power interruptions for up to 2.5 hours. For continuous use over an entire working day (8 hours) at field sites lacking mains power, portable diesel generators were predominately used. However, voltage fluctuations from these generators reduced the reliability and lifespan of the UPS in off-grid environments. Lithium power bank batteries were later found to be a more affordable alternative, offering longer operation times (up to 3.5 hours on a 20,000 mAh capacity) with significantly smaller volume and weight, and the ability to be charged simultaneously from common power sources including USB-C chargers or solar panels. Nevertheless, these batteries presented restrictions during air transport for field trials, as they are permitted only in carry-on luggage and not in checked baggage. Recommendations to address these challenges include incorporating voltage stabilisers or more robust power regulation to mitigate fluctuations, standardising around power banks for power input, clarifying air transport guidance, enabling hot swapping of power banks, and testing solar recharging options.

#### 3.2.3 Scanning Performance

Users found the equipment setup straightforward, enabling them to assemble and initiate slide scanning in under 30 minutes with minimal guidance, training, or instructions. However, initial scan times of 25–35 minutes per slide were deemed too slow for efficient fieldwork, although later improvements reduced this to an average scan time of 12.5 (±6 SD) minutes per slide, with variability due to sample preparation and sample area with 622 (±102 SD) fields of view images per scan. Despite users being able to parallelise scanning and perform other tasks during scanning, feedback from both countries emphasised a desire for faster scan times. Additionally, hardware issues were noted, including one scanner becoming inoperable due to failed glue on the magnetic pickup for slides, preventing scanning operation. Recommendations include optimising the scanning algorithm and optics to further reduce scan time, as well as providing maintenance training, repair manuals, and local repair kits to address hardware failures. Other challenges observed during fieldwork, though not directly in **Table 2**, included susceptibility to insects entering WSI scanners during nighttime operations due to attraction to the bright light source, highlighting the need for an automatic closing flap on the slide loading port, and failures of Ethernet cables due to repeated setup and pack-down in outdoor environments, which were later mitigated by a software update enabling wireless image transmission.

#### 3.2.4 Software & Interface

Users initially performed manual annotations of helminth eggs using the EggInspector application on a laptop, a laborious but essential task for developing the database for AI model training. This process was better suited to a laboratory or office environments and was most effective with a mouse due to the precision required to draw bounding boxes around objects; otherwise, users from both sites found the application user-friendly and easy to use. Once the AI model was trained, reviewing AI-detected objects on the laptop became more streamlined, focusing only on the egg-like objects. However, challenges included the lack of a high-contrast mode on the screen, which was problematic when operating in direct sunlight. Additionally, while the touch screens were capable of operation with disposable gloves, switching between tasks such as sample preparation, slide loading, and egg verification required careful consideration to glove disposal and equipment sterilisation. Users noted that verification work could be deferred to a later stage (e.g., after returning from the field) since images and data were preserved. Although laptop battery life was sufficient to sustain a full day of work, verifying eggs in field scenarios was considered potentially more suitable on a mobile device.

#### 3.2.5 Training & Support

Users emphasised the value of practical training over written instructions, finding hands-on training sessions far more effective than paper manuals for mastering the AI-DP platform’s operation. While remote support was useful for troubleshooting, field maintenance remained challenging due to limited on-site resources and technical expertise. Recommendations include expanding training programs with local biomedical support materials and instructional videos, as well as developing a printable or online troubleshooting manual to assist with common issues. Additionally, options for engaging local support technicians should be explored to enhance field support capabilities.

## 4 DISCUSSION

In this study, we present (i) the technical details of the AI-DP platform designed based on lessons learned from field and laboratory evaluations, (ii) the analytical performance of the AI model, and (iii) the user experience in both controlled and real-world settings.

### Advancing NTD Monitoring: AI-DP Enables Near-Real-Time Data with Quality Assurance and Reduces Technician Burden

The AI-DP platform offers a promising approach to M&E of NTDs such as STH and intestinal SCH. By integrating WSI scanners, onboard AI analysis, and human-in-the-loop verification, it provides near-real-time, quality-assured data while reducing the burden on technicians. Iterative design improvements—guided by earlier challenges [18]—included reducing magnification from 10× to 3.2× for faster scanning, adopting high frame rate focus stack acquisition to handle uneven KK smear thicknesses, and optimising hardware for portability (e.g., rugged case under 32 kg, battery-powered operation). Compared to other devices in the field, such as those reviewed [15–17], our platform uniquely combines field-deployable hardware with offline AI inference and EDC, making it suitable for zero-infrastructure settings. Unlike many low-cost optical devices that prioritise cost and simplicity over throughput, our system aims to match or exceed manual microscopy turnaround times, though scan times (currently 10-12.5 minutes per slide) still lag behind expert manual reading (often under 5 minutes per slide). Nevertheless, automation frees technicians for parallel tasks, promising substantial labour savings—critical given that personnel costs constitute 42–74% of M&E survey expenses [9].

### Addressing Ground Truth Data Quality and Class Imbalance: Key Challenges in AI Model Performance

Despite achieving high precision (84.6–95.9%) and recall (79.1–91.7%) across species using the YOLOv8n model, significant challenges persist in our ground-truth database of 43,919 eggs from 951 slides. Class imbalance, particularly for underrepresented species like hookworm (1.2% of eggs) and *S. mansoni* (3.0%), likely contributes to lower recall for *S. mansoni* (79.1%) and precision for hookworm (84.6%). Equally critical is the quality of ground truth labels, as the annotation process involves rigorous and time-consuming verification effort by multiple parasitologists to ensure accuracy. Ideally, triple verification should be achieved on all images to maximise label quality. However, user fatigue during prolonged verification sessions poses an additional challenge, potentially compromising the consistency and quality of annotations. Future efforts will prioritise expanded data collection for underrepresented species, alongside advanced AI techniques such as class weighting, custom data loaders, and data augmentation to address imbalance. Additionally, strategies to streamline verification—such as automated pre-annotation tools or user-friendly interfaces to reduce fatigue—and research into out-of-distribution performance will enhance model generalisability and robustness in diverse field conditions [27], ensuring consistent accuracy across varying sample qualities and preparation methods encountered during real-world deployment.[27]

### Fostering Country Ownership: Co-Developing AI-DP through Local User Feedback for Field Settings

Collaboration with field users across Ethiopia and Uganda has been central to refining the AI-DP platform, emphasising country ownership and co-development to ensure relevance in resource-constrained settings. Feedback from 14 local users over 30 real-world field scenarios in these countries drove hardware improvements (e.g., integrating routers into scanner housing to reduce cable clutter, adding handles for portability) and software enhancements (e.g., over-the-air updates to resolve Ethernet cable failures, improved EggInspector interface with high-contrast mode for sunlight visibility). Engaging local technicians and researchers not only tailored the platform to address specific field challenges but also built capacity and fostered a sense of ownership among national stakeholders. Users noted that scan times remain a bottleneck compared to manual microscopy, prompting planned optimisations like AI-driven focus prediction to reduce Z-stack acquisition. Future developments, shaped by ongoing dialogue with local partners, will include a mobile verification app for field convenience, streamlined QR printing workflows, and controlled manufacturing processes to ensure durability under harsh conditions. These iterative changes, grounded in a co-development approach, underscore the platform’s adaptability to the unique needs of endemic regions and its potential for sustainable integration into national health systems.

### Validating Performance: The Critical Next Step for AI-DP Adoption

While prior fieldwork in Peru demonstrated the AI-DP platform’s higher sensitivity at low infection intensities compared to manual microscopy [28], comprehensive validation is essential to establish its role in large-scale deworming programs. A detailed evaluation protocol [19] is underway in Ethiopia and Uganda, targeting school-age children to assess: (i) diagnostic performance, (ii) repeatability and reproducibility, (iii) time-to-result, (iv) cost-efficiency, and (v) usability in laboratory and field settings. This non-inferiority study against manual KK screening will quantify efficiency gains and inform scalability. Additionally, integration with national health information systems raises regulatory, data-security, and privacy considerations that must be addressed for sustainable adoption. Sample preparation variability across sites further highlights the need for automated quality-control checks to reject substandard smears before scanning [28][19].

### Exploring Scalability: Potential of AI-DP for Broader NTD Diagnostics

The modular design of the AI-DP platform and its adaptable AI-training framework position it as a versatile tool beyond STH and SCH. Low-hanging fruit includes adapting the system for urinary SCH (*S. haematobium*), where current magnification settings remain suitable, though urine filter samples may present challenges similar to those overcome with KK thick smears, such as variable thickness and the need for multiple focus planes to identify morphological features of eggs. Similarly, diagnostics for lymphatic filariasis using blood thick smears can leverage existing imaging parameters, as these smears are typically more uniform (monoplane) and microfilariae are larger than STH eggs, potentially simplifying detection. Other microscopy-based NTD diagnostics, such as mycetoma and scabies, could be incorporated with adjustments to AI models trained on relevant datasets and, if needed, minor tweaks to imaging protocols and magnification [29]. Beyond human health, the platform holds potential for One Health applications, supporting diagnostics in soil and animal health contexts in resource-limited settings, which could enhance the cost-benefit ratio of the scanner by broadening its utility across sectors. This scalability aligns with WHO’s 2030 NTD elimination goals [1], offering a unified diagnostic platform for resource-limited environments. Future work will explore these expansions, ensuring hardware and software flexibility to meet diverse diagnostic needs across human, animal, and environmental health domains.

## 5 CONCLUSIONS

The AI-DP platform offers a promising approach to field-deployable, AI-enhanced microscopy for NTD M&E programs. By enabling near-real-time, quality-assured data and reducing the workload on technicians, it supports the potential for improved efficiency in STH and intestinal SCH control programs within endemic, resource-constrained settings. Its modular design and adaptable AI-training framework facilitate expansion to other microscopy-based NTD diagnostics, such as urinary schistosomiasis and lymphatic filariasis, as well as broader applications in One Health contexts, contributing to global elimination targets. However, comprehensive validation studies are essential to evaluate its clinical diagnostic performance, repeatability, cost-effectiveness, and usability across diverse field conditions in large-scale deworming programs, ensuring its readiness for sustainable implementation.

## 6 DATA AVAILABILITY

Both the code and image datasets used to develop the AI models are available at Kaggle under a Creative Commons Attribution-ShareAlike 4.0 International (CC BY-SA 4.0) licence:

Data: https://www.kaggle.com/ds/3675429 [DOI: 10.34740/kaggle/ds/3675429]

Code: https://www.kaggle.com/code/mohaliy2016/ai4ntd-kk2-0-p3-0-sth-schm

Correspondence should be addressed to Peter Ward.

## 7 ACKNOWLEDGEMENTS

We thank the Ministry of Health vector-control teams, research teams, field technicians, school administrators, and community leaders in Ethiopia and Uganda for their support in implementing and evaluating the AI-DP platform. We are also grateful to all study participants and their families for their cooperation.

Additionally, we would like to acknowledge all AI4NTD consortium members who contributed to the project. The AI4NTD consortium consisted of Johnson & Johnson (formerly Janssen Pharmaceutica N.V. division of Johnson & Johnson Global Public Health Research & Development; hereafter J&J), Merck KGaA (Merck), Stitching Lygature (Lygature), Bridges to

Development (Bridges), Ghent University (UGent), Jimma University (JU), Ethiopian Public Health Institute (EPHI), Etteplan Sweden AB (Etteplan), Division of Vector Borne and Neglected Tropical Diseases and Ministry of Health, Uganda (VB&NTD). Merck and J&J provided funding, and J&J provided in-kind support on scientific matters.

Etteplan was the work package lead for development and production of the system and technology. Jimma University was the work package lead for the evaluation of the AI-DP platform against TPPs. EPHI and VB&NTD were the work package leads for field evaluation of the AI-DP platform in Ethiopia and Uganda programmatic settings. Bridges was the work package lead for preparing for policy, access and integration into data and country-health systems. Lygature was the consortium coordinator and work package lead for project management and communication. The consortium partners worked across work packages in order to develop and validate the platform in field.

Consortium Members:

- Bridges to Development: Alan Brooks, Julie Jacobson, Anastasia Pantelias, Kayla Hendrickson
- Ethiopian Public Health Institute: Gemechu Leta, Kalkidan Begashaw
- Etteplan Sweden AB: Peter Ward, August Tynong, Joel Larsson, Gustav Jonsson-Glans, Mattias Adolfsson, Anders Kaplan, Hannes F. Kuchelmeister, Maija Persson, Matthias Zumpe, Peter Dahlberg
- Ghent University: Bruno Levecke, Mieke Kennis, Sofie Van Hoecke, Mohammed Ali Mohammed
- Janssen Pharmaceutica NV: Ole Lagatie, Christof Steyaert, Angelo Trotta
- Jimma University: Mio Ayana, Abebaw Tiruneh, Hundaol Girma, Daniel Dana, Zeleke Mekonnen
- Merck KGaA: Mireille Gomes, Béatrice Greco
- Stitching Lygature: Kristina Orrling, Karin de Ruiter
- Division of Vector Borne and Neglected Tropical Diseases and Ministry of Health, Uganda: Betty Nabatte, Semakula Moses, Bogere Ronald, Chris Opio, Narcis Kabatereine

## 8.1 Author Contributions

- Peter K. Ward: Conceptualisation; Methodology; Software; Formal analysis; Visualisation; Writing – original draft.
- Mohammed A. Mohammed: Data curation; Software; Formal analysis; Writing – review & editing.
- Mio Ayana: Investigation; Methodology; Field Supervision; Data curation.
- Lindsay A. Broadfield: Methodology; Supervision; Writing – review & editing.
- Peter Dahlberg: Investigation; Resources; Project administration.
- Daniel Dana: Investigation; Methodology; Field Supervision; Data curation.
- Narcis Kabatereine: Investigation; Resources; Project administration.
- Gemechu Leta: Investigation; Resources; Project administration.
- Zeleke Mekonnen: Investigation; Resources; Project administration.
- Betty Nabatte: Investigation; Methodology; Field Supervision; Data curation.
- Kristina M. Orrling: Project administration.
- Sofie Van Hoecke: Conceptualisation; Methodology.
- Bruno Levecke: Investigation; Methodology; Writing – review & editing.
- Lieven J. Stuyver: Conceptualisation; Supervision; Funding acquisition; Writing – review & editing.

Refer to the **Acknowledgements** for a complete list of AI4NTD consortium members.

## 8.2 Competing Interests

The authors and funders declare the following competing interests:

After completion of the study, IP and technology related to the platform were divested from J&J to Enaiblers to bring the solution to market. Peter Ward and Peter Dahlberg are employed at Enaiblers and own stock/shares in Enaiblers. Lindsay Broadfield and Lieven Stuyver are scientific advisors to Enaiblers.

Merck KGaA drug praziquantel is the standard treatment of care for schistosomiasis. Since 2007, Merck KGaA has donated more than 1.5 billion Praziquantel tablets, enabling the treatment of more than 600 million school-aged children. Merck KGaA has committed itself to maintaining its efforts in the fight against the tropical disease until schistosomiasis has been eliminated. To this end, each year Merck KGaA is donating up to 250 million Praziquantel tablets to the World Health Organization.

LJS was the Scientific Leader of the AI4NTD steering committee and a stockholder of J&J.

